# Gametocyte production and infectivity among Ugandan malaria patients infected with *P. falciparum* with partial resistance to artemisinins

**DOI:** 10.64898/2026.04.09.26350477

**Authors:** Daniel Ayo, Sara Lynn Blanken, Ismail Onyige, Eric Musasizi, Nicholas Proellochs, Thomas Katairo, Shahiid Kiyaga, Bienvenu Nsengimaana, Innocent Wiringlimanna, Francis D. Semakumba, Isaac Ssewanyana, Kjerstin Lanke, Douglas Opiyo, Moses R. Kamya, Arjen Dondorp, Tonny Etwop, Smart G. Okot, Bryan Greenhouse, Liusheng Huang, Jessica Briggs, Laura E. de Vries, Grant Dorsey, Chris Drakeley, Philip J. Rosenthal, Melissa D. Conrad, Emmanuel Arinaitwe, Maurice Okao, Teun Bousema

## Abstract

**Background:** Partial resistance to artemisinins (ART-R) has emerged in East Africa, associated with mutations in the *Plasmodium falciparum kelch13* gene. It is currently unclear whether ART-R has implications for gametocyte production or for onward transmission to mosquitoes.

**Methods:** In a cohort of uncomplicated malaria patients attending Kalongo Hospital in northern Uganda, we quantified carriage of PfKelch13 mutant parasites by conventional sequencing and droplet digital PCR (ddPCR) for the C469Y and A675V mutations. Prevalence and density of gametocytes and ring-stage parasites were assessed by microscopy and quantitative reverse-transcriptase PCR (qRT-PCR). Lumefantrine concentrations, indicative of prior malaria treatment, were determined by ultra-high performance liquid chromatography-tandem mass spectrometry. Transmission potential of wild-type and PfKelch13 mutant parasites was assessed by mosquito feeding assays and complemented with molecular characterization of parasites in wild-caught mosquitoes from household resting catches.

**Findings:** We enrolled 235 patients with symptomatic *P. falciparum* infection; PfKelch13 C469Y or A675V mutations were detected in 35.8% (78/218) of infections by sequencing and 59.1% (136/230) by ddPCR. Gametocyte carriage was 24.0% (56/233) by microscopy and 56.6% (133/235) by qRT-PCR and not associated with the abundance of PfKelch13 mutant parasites by ddPCR (p=0.603). Among a total of 227 mosquito feeds with patient whole blood, 1.4% (120/8745) of mosquitoes became infected. Mosquito infection rates were positively associated with gametocyte density (β = 0.39, 95% CI = 0.23-0.59, *p* < 0.001) without an observed interaction with the abundance of PfKelch13 mutant parasites (p = 0.452). PfKelch13 C469Y or A675V mutations were detected in 40.1% (21/52) of malaria-infected bloodmeals of field-caught mosquitoes and in 28.0% (7/25) of sporozoite-positive mosquitoes.

**Interpretation:** We conclude that *pfkelch13* mutations are very common in patients in northern Uganda with uncomplicated malaria, mostly in multiclonal infections. We observed no evidence that ART-R affected gametocyte production or transmission to mosquitoes.

**Funding:** Dutch Research Council (NWO)

**Research in context:** *Evidence before this study:* Partial resistance to artemisinins (ART-R) might enhance or inhibit gametocyte formation or transmission of malaria parasites to *Anopheles* mosquitoes. However, few studies directly assessed *P. falciparum* transmission potential in relation to sensitivity to ART-R or associated mutations. We searched PubMed on January 14th 2026, with no restrictions on publication date or language, for studies assessing gametocyte carriage or transmissibility to mosquitoes in uncomplicated malaria cases in relation to artemisinin resistance using the search terms (“uncomplicated malaria” OR “patient”) AND (“gametocyte” OR “anopheles”) AND ((“artemisinin” AND “resistance”) OR “pfkelch13” OR “kelch13”). From the 101 identified articles, the majority did not report on parasite resistance or resistance markers. Nineteen articles reported original patient data, with four additional studies examining parasite isolates for gametocyte production and transmissibility *in vitro*. *In vitro* studies all reported gametocyte formation in parasite isolates with *pfkelch13* mutations and, for three studies where transmission was directly determined, evidence for successful mosquito infections from ART-R parasite isolates. None of these studies demonstrated consistent differences in gametocyte production or transmissibility between ART-R and sensitive parasite isolates. One clinical study from Thailand observed higher levels of gametocyte carriage during follow-up for infections with slow asexual blood-stage parasite clearance following artemisinin-combination therapy (ACT) and an increase in gametocyte carriage at clinical presentation over a period when the prevalence of ART-R was rising. A study with patient data from 7 Asian and 3 African countries reported higher proportions of pretreatment and post-treatment gametocytemia in patients with slow parasite clearance. Another study from Cambodia reported a higher prevalence of gametocytes at enrolment in areas affected by ART-R, but no association between gametocyte carriage and either individual-level parasite clearance or treatment failure. In contrast, a meta-analysis of ACT treatment efficacy in pregnant women observed no association between gametocyte carriage at baseline and treatment failure. Only one study, directly examined gametocyte carriage at clinical presentation with *pfkelch13* mutations and found no association between gametocyte carriage and molecular markers of ART-R or *ex vivo* drug sensitivity in Cambodia.

*Added value of this study:* We performed a direct assessment of gametocyte production, carriage of mature gametocytes and transmission to *Anopheles gambiae* s.s. mosquitoes in relation to validated ART-R markers in patients presenting with uncomplicated *P. falciparum* malaria in an area affected by ART-R in Uganda. Our work demonstrates that parasites with the PfKelch13 C469Y and A675V mutations were very common, mostly in multiclonal infections. Gametocyte carriage and gametocyte commitment were similar between patients presenting with pure or predominantly wild-type infections and those presenting with PfKelch13 mutant infections. In feeding studies, mosquito infection rates were similar for patients with wild-type or PfKelch13 mutant infections; wild-caught mosquitoes confirmed ongoing transmission of parasites with molecular signatures of ART-R.

*Implications of all the available evidence:* Taken together, there are no indications for altered gametocyte production or transmissibility to mosquitoes of infections with PfKelch13 C469Y and A675V. Future studies may examine impacts of other mutations in *pfkelch13*, study transmission potential in low endemic settings where monoclonal infections dominate and aim to understand how gametocyte clearance and post-treatment transmission potential may differ between ART-R and wild-type infections upon treatment.

## INTRODUCTION

Prompt treatment with artemisinin-combination therapy can prevent severe consequences of malaria and reduce transmission to mosquitoes. Partial resistance to artemisinins (ART-R) manifests as delayed parasite clearance upon ACT treatment and is associated with mutations in the gene sequence corresponding to the propeller domain of the *Plasmodium falciparum* Pfkelch13 protein (1). ART-R first emerged in Southeast Asia (2), and more recently in East Africa (3–5). Validated markers of ART-R in East Africa include A675V, C469Y and R561H (4), of which the first two are highly prevalent in northern Uganda (6).

The fate of ART-R parasites is determined by their ability to survive in human populations and to spread through mosquitoes. Mutations that confer ART-R may incur a substantial fitness cost during asexual parasite multiplication(7); for some mutations this results in a lower ability to survive in competition with other clones(7, 8) and a lower prevalence in naturally acquired multiclonal infections (9). Sensitive quantitative techniques, including droplet digital PCR (ddPCR), may improve the detection and quantification of ART-R parasites in multiclonal infections compared to conventional Sanger sequencing (10). Upon ingestion by mosquitoes, mature gametocytes activate and fertilize to form oocysts on the midgut wall that grow over time and release sporozoites that render the mosquito infectious to humans. ART-R parasites may experience a different transmission potential compared to wild-type parasites. Gametocyte prevalence at clinical presentation is higher among patients who subsequently experience slow parasite clearance following artesunate treatment (11) and in areas with high levels of ART-R (12). This suggests higher (intrinsic) gametocyte production for ART-R parasites that is potentially explained by upregulation of AP2-G3 transcription, linked to the master regulator of gametocyte production AP2-G (13), although a study in Cambodian patients did not observe an association between validated markers of ART-R and microscopy-detected gametocyte carriage (14).

Understanding the impact of ART-R on transmission is highly relevant to inform strategies to reduce the spread of this resistance. Here, we quantified gametocyte carriage and gametocyte commitment markers in relation to the presence of *pfkelch13* mutations, duration of symptoms, and prior treatment for malaria in patients seeking care for uncomplicated malaria in northern Uganda. We determined human-to-mosquito transmission by mosquito feeding assays and assessed parasite prevalence in wild-caught mosquitoes, deploying ddPCR for sensitive quantification of common *pfkelch13* mutations.

## METHODS

### Study Site

We conducted two complementary studies in Agago District in northen Uganda, an area with intense malaria transmission. In the first study, we quantified ART-R parasite carriage and transmission potential among uncomplicated malaria cases attending Kalongo Hospital. Patients or their caretakers provided written informed consent; assent was provided for children aged 8-17 years. In the second study, we sampled mosquitoes in households in Kalongo and neighboring villages to assess carriage of ART-R parasites. For mosquito sampling, heads of households provided consent. Our data collection was part of a broader study on the impact of parasite and mosquito factors on malaria transmission in Uganda that received ethical approval from the Uganda National Council for Science and Technology, Makerere University School of Medicine, the University of California San Francisco, and the London School of Hygiene and Tropical Medicine.

### Patient sample collection, plasma drug level assessments and mosquito membrane feeding assays

We recruited patients aged ≥2 years with uncomplicated malaria, defined as the presence of *P. falciparum* mono-infection by microscopy or rapid diagnostic test (RDT), with fever or reported fever in the last 24 hours and no danger signs of severe disease. Patients donated a single blood sample and subsequently received treatment with artemether-lumefantrine according to national guidelines. Blood was collected prior to treatment in heparin tubes (Lithium Heparin; BD Biosciences, Plymouth, UK) for mosquito feeding assays and plasma drug levels and EDTA tubes (Greiner-Bio-K 3 EDTA) for molecular assays. Lumefantrine levels, indicative of prior antimalarial drug exposure, were assessed in plasma samples from 61 patients with *pfkelch13* mutant infections and 85 randomly selected samples with pure wild-type infections using an ultra-high performance liquid chromatography-tandem mass spectrometry system [Waters UPLC (I class) coupled with a Sciex 6500+ triple quad mass spectrometer, equipped with a Zorbax Eclipse Plus C_8_ column (2.1 x 50 mm, 1.8 μm)] (15). This was validated across a calibration range of 50-20,000 ng/mL, with a lower limit of quantification of 50 ng/mL(15). To identify lumefantrine levels consistent with AL treatment in the previous 21 days, we used the 97.5^th^ percentile of a previously described pharmacokinetic model (16) incorporating the age distribiton of our cohort, resulting in a threshold of 170 ng/mL.

Lithium-heparin blood was offered to approximately 60–80 *Anopheles gambiae* s.s. mosquitoes through membrane feeding assays at physiological concentration (‘whole blood’) or after 6 to 14-fold enrichment of gametocytes by magnetic activated cell sorting (‘enriched’), as described previously(17). For each condition, two cups of mosquitoes were placed under two temperature-controlled glass feeders that each contained 0.35-0.5 mL of blood (18). Fully fed mosquitoes were maintained on 10% glucose for 8 days at 25 ± 2^0^C and 80 ± 10% humidity until dissection at day 9-11 post blood feed in 1% mercurochrome and examination for oocysts by two independent microscopists. Infected midguts were transferred to oocyst lysis buffer (18) for molecular analyses.

### Mosquito collections

Indoor resting mosquito sampling was done from January - April, 2024 in 10 villages in Agago District using Prokopack aspirators (Model 1419; John W. Hock Company, Gainesville, Florida, USA) (19). Resting mosquitoes were collected between 4 and 8 am from walls and ceilings of households of study participants and neighbouring households. Households were selected based on convenience sampling, with prioritization of traditional houses for high mosquito yields. For all *Anopheles* mosquitoes, speciation was done based on morphological keys to differentiate *An. funestus* s.l. from *An. gambiae* s.l. For all blood-fed mosquitoes, including non-vectors (*Culex*), the abdomen was squeezed onto a filter paper (Lasec; Whatman 903^TM^ -IVD) and stored with self-indicating silicagel (6-20 MESH blue; Loba Chemistry Ltd.) at -20°C. All female field-caught *Anopheles* mosquitoes, including those without a bloodmeal, were dissected to separate the head and thorax from the abdomen. Head and thoraces were stored in individual tubes with silicagel at -20°C until further processing for *P. falciparum* sporozoite detection and molecular characterization.

### Nucleic acid extraction

There were four sources of material for molecular analyses: i) human patient blood; ii) midguts of mosquitoes that became infected following membrane feeding; iii) bloodmeals of wild-caught mosquitoes; iv) head and thoraces of wild-caught mosquitoes. Human blood aliquots (100 μL EDTA blood) in RNAprotect were subjected to total nucleic acid extraction using the MagNAPure MP96 automated extractor (Roche, Basel, Switzerland) and the MagNAPure 96 DNA and Viral NA Large Volume kit. Head and thorax samples from field-caught mosquitoes were homogenized by mechanical plate shaking using 1mm zirconia beads (Biospec) in a 96-deepwell plate (20). This mosquito homogenate and mosquito midguts from membrane feeding experiments were incubated overnight at 56°C with Proteinase K (Qiagen, RP103B). Mosquito bloodmeals from wild-caught mosquitoes were first treated with Proteinase K diluted in ATL buffer (Qiagen, 939011); incubated overnight at 56°C and subsequently boiled at 95°C for 5 minutes, after which MagNA Pure 96 External Lysis buffer (Roche, 06374913001) was added. All mosquito materials were subsequently used for automated extraction of nucleic acids using the MagNAPure DNA and Viral NA Small Volume kit.

### Molecular analyses on human blood samples and mosquito materials

Extracted nucleic acids (NA) from human blood was used for quantitative reverse transcription PCR (qRT-PCR) to quantify asexual blood-stage parasite density based on ring-stage *sbp1* transcripts (21), gametocyte commitment based on *ap2-g* transcripts (21) and mature gametocyte density based on male (*pfmget*) and female (*ccp4*) gametocyte transcripts (22). qRT-PCR gametocyte prevalence and density were adjusted for low levels of gametocyte transcripts from asexual blood-stage parasites (23); for patients who were microscopy positive for gametocytes but with negative measures of PfMGET-CCp4, microscopy gametocyte density was used. The same NA material was used for Sanger sequencing the *pfkelch13* propeller domain (codons 440–680) following PCR purification (QI-quick PCR Purification Kit)), and for amplicon sequencing using the MAD^4^HaTeR panel (Paragon Genomics, catalog PGD268) with a multiplex PCR-based CleanPlex workflow to estimate complexity of infection (COI) using MOIRE (24), which was calculated as the maximum number of alleles observed across all genotyped loci, accounting for allele frequencies and within-host relatedness. Lastly, a multiplex ddPCR assay was developed to sensitively detect and quantify C469Y and A675V mutations. Total NA samples were diluted based on parasite density estimates to prevent droplet overload. Droplets were generated with the QX200 Automated Droplet Generator, moved to the C1000 Touch Thermal Cycler for thermal cycling (56°C annealing), followed by readout on the QX200 Droplet Reader and analysis by QuantaSoft or QX Manager software. Each reaction analyzed a minimum of 10,000 droplets (range 10,000-22,000 droplets) to enable correct absolute quantification of mutant and wild-type alleles. In case a reaction had less than 100 negative droplets, the sample was diluted and repeated to avoid incorrect quantification due to droplet overload. Since A675V mutant parasites gave a wild-type signal in the multiplex ddPCR (**Supplemental Methods**), A675V positive samples were rerun in the single A675V SNP assay for correct quantification. Of note, ddPCR quantified C469Y and A675V mutations only; parasite populations without these two mutations were classified as wild type.

Extracted nucleic acids from mosquito material (midguts from membrane feedings; bloodmeals or head-thorax homogenates from wild-caught mosquitoes) were first tested by quantitative PCR (18); positive samples with a CT-value below 35 were processed by ddPCR as described above. Details on molecular methods and number of successful assays are provided in **Supplemental Methods and Results**.

### Statistical analysis

The sample size for enrolled malaria patients was based on an estimated 30% prevalence of PfKelch13 mutant infections (4), an estimated qRT-PCR gametocyte prevalence of 35% in uncomplicated malaria cases with wild-type parasites (25) and a hypothesized ≥75% higher gametocyte prevalence in mutant infections (i.e. prevalence of 61.3%) (26). Enrolling 230 patients (of whom ∼69 would have PfKelch13 mutant infections) would give >99% power to detect a difference in gametocyte prevalence with a one-sided alpha of 0.05. In the absence of prior data on sporozoite rates in the study area, we performed no sample size calculations for the household collection of resting mosquitoes. Total parasite and gametocyte densities were log transformed and modelled using a Gaussian distribution. Linear regression models were used to determine the association between gametocyte and ring-stage parasite densities and between gametocyte density and mosquito infectiousness (25). Models were run in the presence and absence of an interaction between gametocyte density and the proportion of PfKelch13 mutant parasites (in categories, <10, 10-49, ≥50%), to assess whether the association between gametocyte density and mosquito infectivity differed based on the proportion of PfKelch13 mutants. Associations between numerical variables were assessed using Spearman’s rank correlation, Kruskal-Wallis tests were used to assess differences in numerical variables between groups, and two-sample Z-tests were used to assess differences in proportions.

## RESULTS

Between May 2023 and June 2024, we enrolled 235 patients with uncomplicated malaria. Infection with *P. falciparum* was confirmed by microscopy in 99.6% (234/235) of patients; one patient was positive by Rapid Diagnostic Test, negative by microscopy, but *P. falciparum* positive by molecular methods. The median age of patients was 13 years (IQR: 6–19), and median haemoglobin concentration 11.4 g/dL (IQR: 9.9–12.8). Most patients (70.2% (165/235)) were afebrile at presentation but reported symptoms in the past 24 hours (**Table 1**). Median asexual blood-stage parasite density by microscopy was 13,880 parasites/µL (IQR: 1,260–117,960) in children aged 2–4 years (n=51), 19,520 parasites/µL (IQR: 5,780– 94,140) in children aged 5–15 years (n=87), and 14,140 parasites/µL (IQR: 2,290– 54,850) in older individuals (n=96) (**Supplementary Figure 2A**; Kruskal–Wallis test: p=0.27). Asexual blood-stage parasite densities by microscopy were positively correlated with the ring-stage parasite burden quantified by *sbp1* qRT-PCR (Spearman’s ρ=0.42, *p* < 0.0001) **(Supplementary Figure 2B).** Among the 215 infections with successful COI estimates, 13% (28/215) were monoclonal and 87% (187/215) were multiclonal, with a median of 3 clones per infection (IQR 2-4; maximum of 14 clones). Microscopy gametocyte prevalence was 23.1% (12/52) in children <5 years, 24.4% (21/86) in children aged 5–15 years, and 24.2% (23/95) in older individuals. Including qRT-PCR, gametocyte prevalences were 57.7% (30/52), 55.2% (48/87), and 57.3% (55/96), respectively (**Table 1**). Lumefantrine was detected in 46.2% (72/156) of plasma samples (>50 ng/mL) with a wide range of estimated drug levels (**Supplementary Figure 3**, **Table 1**).

**Table 1:** General characteristics of study patients. Current fever was measured at a temperature of ≥ 37.5°C, a history of fever in the past 24 hours and the duration of malaria-related symptoms was self-reported. Ring-stage parasite (qRT-PCR, sbp1) and gametocyte (qRT-PCR, PfMGET, CCp4) prevalence and densities (amongst positive observations) are indicated per age category. Complexity of infection (COI) estimates were used to distinguish monoclonal from polyclonal infections; the median number of clones was calculated for polyclonal infections. Lumefantrine prevalence was calculated for all positive plasma levels (>0 ng/mL), and for observations with plasma concentrations consistent with artemether lumefantrine treatment in the previous 21 days (> 170 ng/mL).

|  | Total | Age category |  |  |
| --- | --- | --- | --- | --- |
|  |  | < 5 years | 5 – 15 years | > 15 years |
| Current fever<br>(%, n/N) | 28.9<br>(68/235) | 38.5<br>(20/52) | 37.9<br>(33/87) | 15.6<br>(15/96) |
| History of fever (24 hours)<br>(%, n/N) | 99.6<br>(233/234) | 100.0<br>(52/52) | 98.9<br>(86/87) | 100.0<br>(95/95) |
| Duration of symptoms<br>(hours, median, IQR) | 72 (54-102) | 78 (48-102) | 60 (54-104) | 72 (55-96) |
| Ring-stage parasite<br>prevalence by qRT-PCR<br>(%, n/N) | 97.9<br>(230/235) | 98.1<br>(51/52) | 100.0<br>(87/87) | 95.9<br>(92/96) |
| Ring-stage parasite density<br>by qRT-PCR<br>(per $\mu\text{L}$, median, IQR) | 3851.0<br>(478.7-15601.2) | 5515.2<br>(455.0 – 32343.6) | 6935.7<br>(1030.8-15921.3) | 2453.7<br>(237.8-6941.4) |
| Gametocyte prevalence by<br>qRT-PCR<br>(%, n/N) | 56.6<br>(133/235) | 57.7<br>(30/52) | 55.2<br>(48/87) | 57.3<br>(55/96) |
| Gametocyte density by<br>qPCR<br>(per $\mu\text{L}$, median, IQR) | 14.5<br>(0.44-81.2) | 66.9<br>(5.4 – 196.2) | 32.0<br>(0.28 – 105.9) | 3.7<br>(0.34-19.1) |
| Polyclonal infection<br>prevalence (%, n/N) | 87.0<br>(187/215) | 87.5<br>(42/48) | 88.6<br>(70/79) | 85.2<br>(75/88) |
| Complexity of infection<br>(median clones, IQR) | 3 (2-4) | 3 (2-4) | 3 (2-4) | 3 (2-4) |
| Lumefantrine prevalence<br>($> 0$ ng/mL) | 46.2<br>(72/156) | 36.4<br>(12/33) | 53.1<br>(34/64) | 44.1<br>(26/59) |
| Lumefantrine prevalence<br>($> 170\text{ng/mL}$) | 10.9<br>(17/156) | 9.1<br>(3/33) | 12.5<br>(8/64) | 10.2<br>(6/59) |

Conventional sequencing of the propeller domain of *pfkelch13* (codons 440–680) was successful in 92.8% (218/235) of samples (success rates for molecular assays are presented in the **Supplemental Results**). The majority (64.2%; 140/218) of samples were classified as pure wild-type infections; mutant alleles were detected in 35.8% (78/218) of samples, predominantly C469Y (29.8%; 65/218) or A675V (3.7%; 8/218). Five samples (2.3%) showed evidence of other PfKelch13 mutations, notably P441A (n=3 of which 1 also had C469Y), E433D (n=1), and R561H (n=1, mixed with wild-type) (**Figure 1A**). Among the 230 infections with ddPCR results, 34.3% (79/230) were pure wild type (at position 469 and 675) and 59.1% (136/230) mixed wild type/mutant (C469Y + A675V, **Figure 1B),** with a wide range in the proportion of gene copies that was mutant (0.008-99.1%) (**Figure 1C**). The likelihood of harbouring any C469Y and/or A675V parasites increased with increasing COI (**Figure 1D**). Fifteen samples were pure *pfkelch13* mutant by ddPCR (6.52%; 15/230), with 9 pure C469Y, 2 pure A675V and 4 harbouring both C469Y and A675V without wild-type parasites. Given the abundance of mixed wild-type-mutant infections, we categorized samples as having no or few mutant alleles (<10% mutant; 60.0%, 138/230), intermediate mutant allele proportions (10–49% mutant; 18.7%, 43/230), or predominantly mutant infections (≥50% mutant; 21.3%, 49/230).

**Figure 1.**
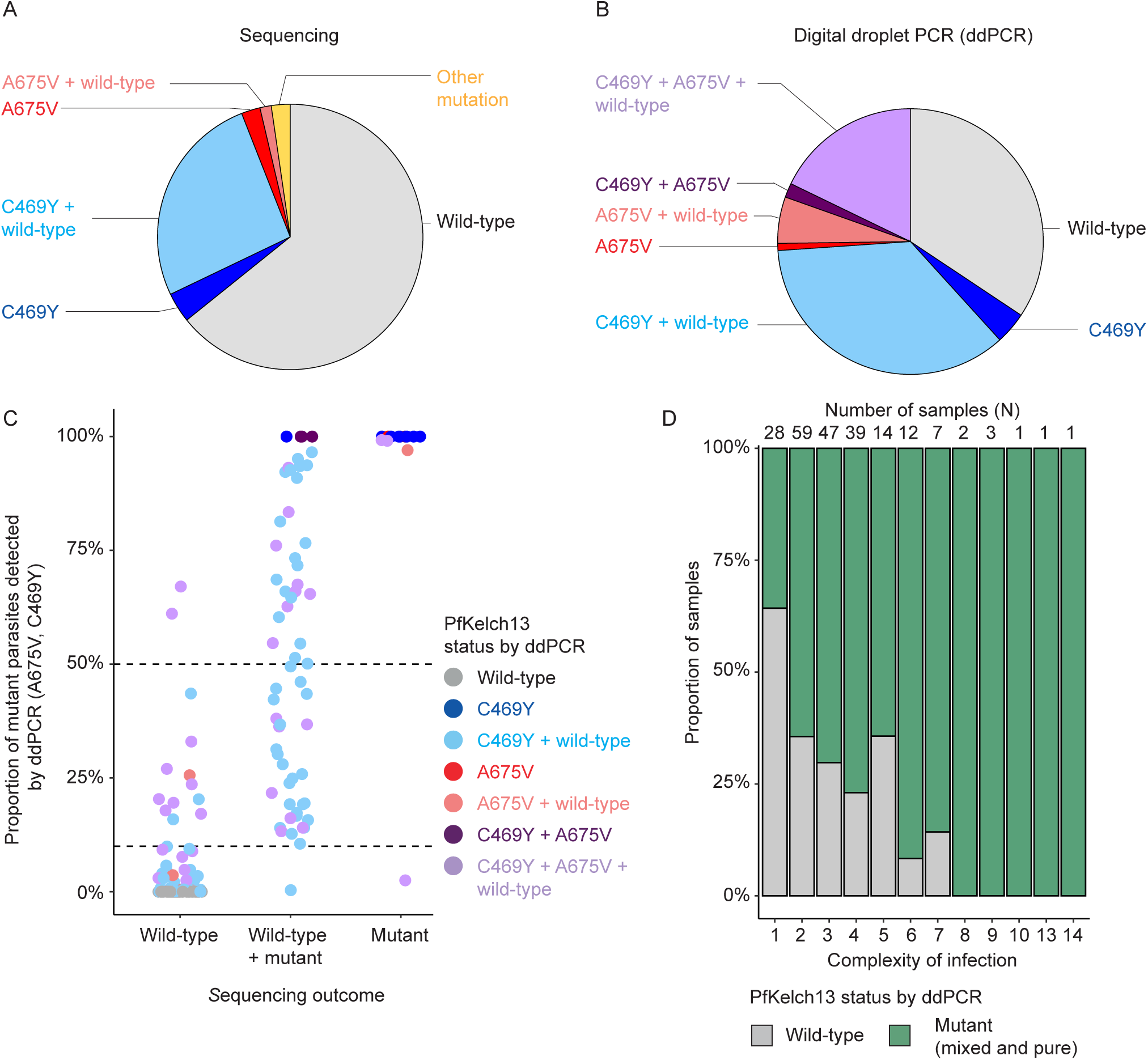
Prevalence and quantity of PfKelch13 mutants in patient whole-blood samples. **AB)** Pie charts showing the proportion of whole-blood samples with PfKelch13 wild-type and mutant parasites determined by **(A)** conventional sequencing and **(B)** digital droplet PCR (ddPCR). Sample numbers per category for sequencing (total N = 218): pure wild-type n = 140; pure C469Y n = 8; mixed C469Y n = 57; pure A675V n = 5; mixed A675V n = 3; other (non-C469Y/A675V) PfKelch13 mutants n = 5. For ddPCR (total N = 230), sample numbers per category were as follows: pure wild-type n = 79; pure C469Y n = 9; mixed C469Y n = 82; pure A675V n = 2; mixed A675V n = 13; pure C469Y and A675V n = 4; C469Y and A675V mixed with wild type n = 41. **C)** Dot plot showing the proportion of PfKelch13 mutant parasites determined by ddPCR in relation to the PfKelch13 mutant status determined by sequencing. The proportion of mutant parasites (C469Y and A675V taken together) is indicated on the y-axis, with dashed lines indicating a proportion of 50% and 10%. On the x-axis, the PfKelch13 mutant status determined by sequencing is indicated in categories. Colours indicate the PfKelch13 mutant categories determined by ddPCR. **D)** Bar chart showing the proportion of wild-type versus mutant samples (determined by ddPCR) in relation to the complexity of infection (in categories). The x-axis indicates the number of clones present in the sample, the y-axis indicates the proportion of samples in that category that have either a pure wild-type or mutant (pure and mixed with wild-type) infection. The number of samples per complexity of infection category is indicated on top.

We had lumefantrine measurements for 83 samples with <10% mutant, 31 samples with 10-49% mutant, and 42 samples with ≥50% mutant parasites. We observed no compelling evidence for a higher prevalence of detectable lumefantrine plasma levels among patients presenting with *pfkelch13*-mutant infections, compared to wild-type infections (p=0.49). Also, when we compared patients with high lumefantrine levels (>170 ng/mL; n=17) with those without detectable lumefantrine, there was no apparent difference in the abundance of PfKelch13 mutant parasites (**Supplementary Figure 4**; Wilcoxon rank-sum test, p=0.701). Parasite density by *sbp1* qRT-PCR was broadly similar between patients with varying proportions of PfKelch13 mutant parasites (**Figure 2A**); a finding that was corroborated by microscopy-based parasite density estimates (**Supplemental Figure 5**). Gametocyte prevalence by microscopy was 26.3% (36/137), 19.0% (8/42), and 22.4% (11/49) in patients with <10%, 10-49% and ≥50% PfKelch13*-*mutant parasites (X^2^=1.01, p=0.603); by molecular methods these prevalences were 55.1% (76/138), 69.8% (30/43) and 53.1% (26/49), respectively (X^2^ = 0.86, p=0.649). We observed no evidence that gametocyte density was associated with the abundance of PfKelch13mutant parasites (**Figure 2B**; Kruskal-Wallis p=0.228). We observed a weak negative association between gametocyte density and the number of ring-stage parasites (β= -0.19, 95%CI = - 0.35 -0.02, p=0.0227) that was not influenced by PfKelch13 mutation abundance (**Figure 2C**). Gametocyte commitment, quantified as the proportion of *ap2-g* over *sbp1* transcripts, was also not associated with PfKelch13 mutation abundance (Kruskal-Wallis p =0.259, **Figure 2D**).

**Figure 2.**
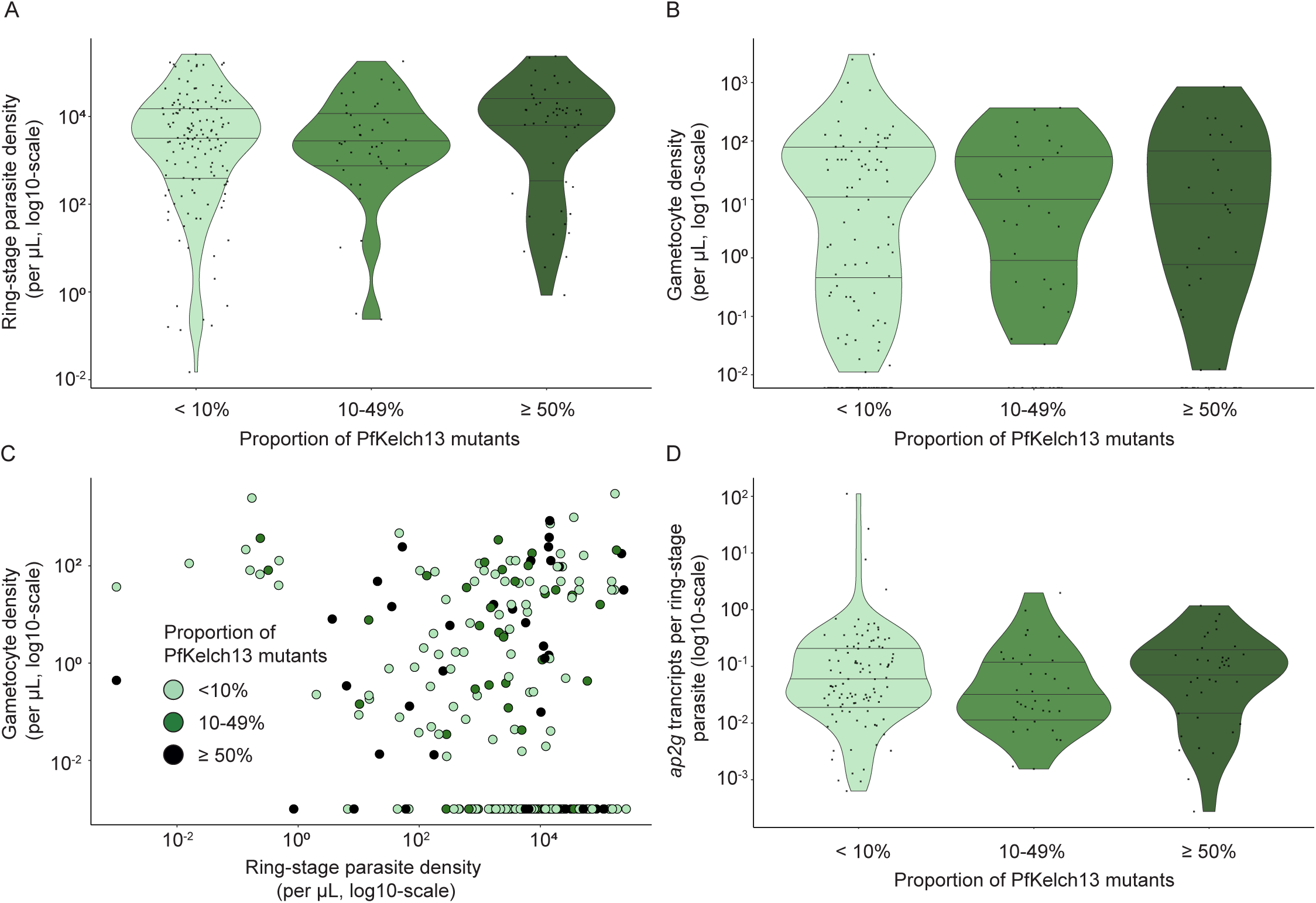
Parasite and gametocyte density in relation to PfKelch13 mutant status. **A)** Violin plot showing ring-stage parasite densities measured in patient whole-blood by qRT-PCR for different categories of proportion PfKelch13 mutant parasites. Samples were divided into three categories based on the proportion of PfKelch13 mutant (C469Y and A675V) infections detected by digital droplet PCR (<10 %, 10-49%, and ≥ 50% mutant parasites). **B)** Violin plot showing gametocyte densities measured in patient whole-blood by qRT-PCR for the three different categories of proportion PfKelch13 mutant parasites. **C)** Scatter plot showing ring-stage parasite densities in relation to gametocyte densities for the different categories of proportion PfKelch13 mutant parasites. Colours indicate the three different proportion of mutant categories. **D)** Violin plot showing the ratio of *ap2-g* transcripts to ring-stage parasite density for the three different proportion of mutant categories.

We conducted mosquito feeding assays on 227 patient whole blood or concentrated gametocyte samples. With whole blood, 9.7% (22/227) of samples infected at least one mosquito and 1.4% (120/8745) of mosquitoes became infected; after gametocyte enrichment 15.4% (35/227) of experiments resulted in at least one infected mosquito and 2.4% (205/8556) of mosquitoes became infected. Gametocyte density was positively associated with mosquito infection rates (**Figure 3A**, **3B)** (whole blood: β=0.39, 95%CI=0.23-0.59, p<0.001; gametocyte-enriched: β=0.32, 95%CI=0.19-0.48, *p*<0.001) without evidence for an interaction with the proportion of PfKelch13 mutant parasites (p=0.452 for whole blood). We successfully genotyped infections in 88 mosquitoes that became infected from whole blood and 144 after gametocyte enrichment. Pure wild-type infections were detected in 76.7% (178/232) of infected mosquitoes; C469Y and A675V in 18.1% (42/232) and 5.2% (12/232) of infected mosquitoes, respectively (Supplementary Figure 6). Samples from six patients exclusively infected mosquitoes with mutant parasites; five infected mosquitoes with both wild-type and mutant parasites (**Figure 3C).** Gametocyte density in whole-blood was positively associated with infection burden in mosquitoes (oocyst density) (β=0.64, 95%CI=0.33-1.02, p<0.001), without an effect of PfKelch13 mutations (**Figure 3D**).

**Figure 3.**
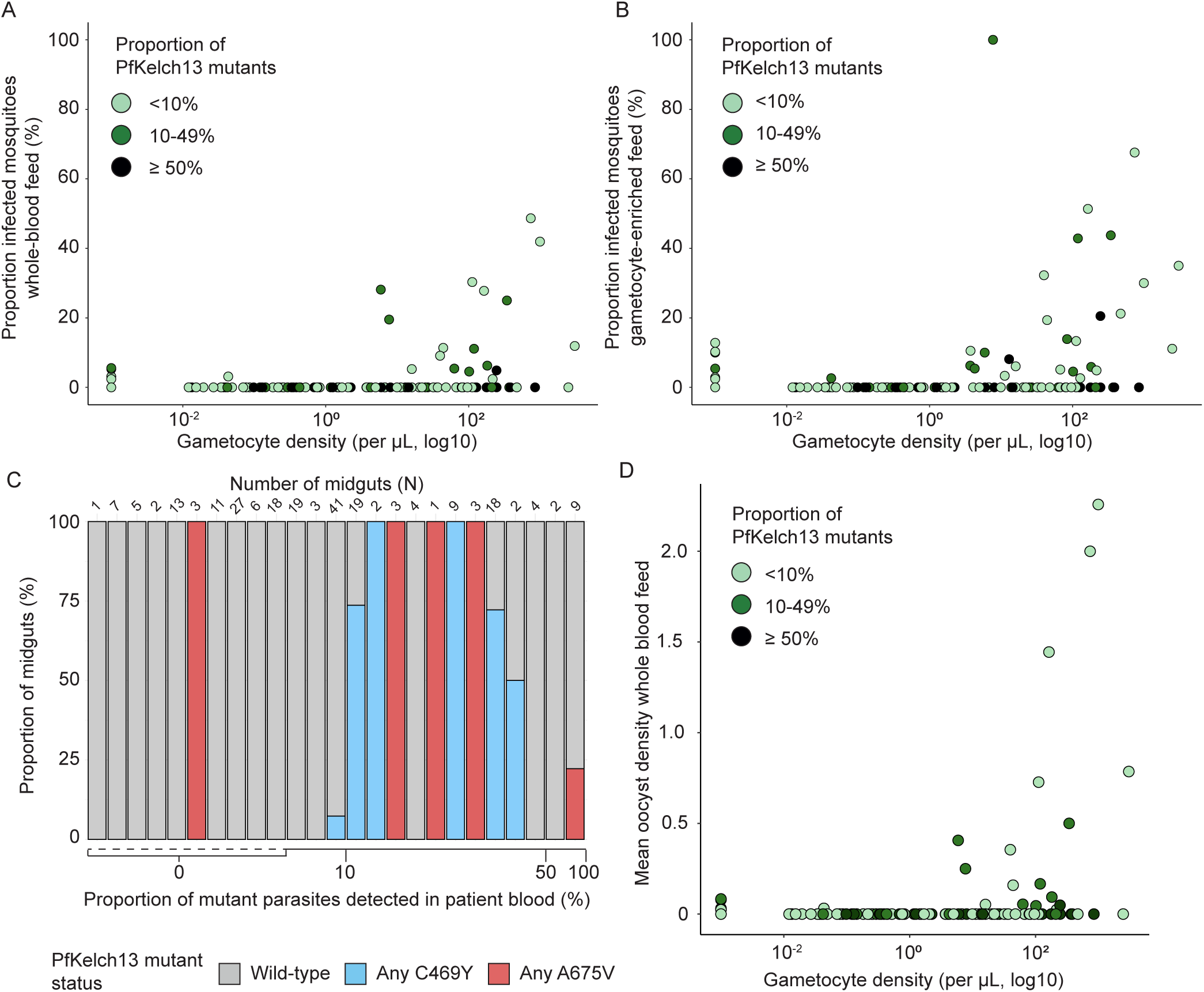
Infectivity of PfKelch13 mutant parasites. **AB)** Scatter plot showing gametocyte densities in relation to the proportion infected mosquitoes for whole-blood **(A)** and gametocyte-enriched **(B)** mosquito feeding assays. Samples were categorized into three groups based on the proportion of PfKelch13 (C469Y and A675V) mutant parasites detected by digital droplet PCR (ddPCR, <10 %, 10-49%, and ≥ 50% mutant parasites). **C)** A stacked bar chart displaying the proportion of mutant parasites detected in patient whole-blood in relation to the proportion of mutant parasites detected in infected mosquitoes from infectious feeding experiments. Every bar displays an infectious individual, combining feeding outcomes of both whole-blood and gametocyte-enriched assays. The proportion of midguts is indicated on the y-axis, the bar colours indicate the PfKelch13 mutant status of infected midguts determined by ddPCR: pure wild-type indicated in grey, pure/mixed C469Y indicated in blue, and pure/mixed A675V indicated in pink. The x-axis depicts the proportion of PfKelch13 mutant parasites (determined by ddPCR) detected in patient whole-blood used for the feeding experiments. The x-axis is non-linear, the dashed line indicates the wide-range of whole-blood samples with zero PfKelch13 mutant parasites detected. Ticks on the x-axis are indicative of the thresholds used for classifying the proportion of PfKelch13 mutant parasites into groups of little (<10%), intermediate (10-49%) and predominantly (≥ 50%) mutant parasites. **D)** Scatter plot showing the relation between gametocyte density and mean oocyst density. Mean oocyst density was calculated per whole-blood feed using all mosquitoes (including non-infected mosquitoes). Samples were again divided into three categories based on the proportion of PfKelch13 mutant parasites detected by ddPCR, which is indicated by the different colours.

Across 395 sampling nights in 200 households (1-4 nights per household), 893 resting mosquitoes were collected. Of these, 22.8% (208/893) were morphologically identified as *Culex spp*., 37.9% (337/893) *An. funestus* s.s, 39.2% (347/893) *An. gambiae s.l.,* and 0.1% (1/893) *An. Zeimanni.* 30.1% (269/893) of mosquitoes, including non-anophelines, were blood-fed and processed for parasite genotyping in blood meals. Among these, *P. falciparum* was detected by 18S qPCR in 39.4% (106/269) of bloodmeals (**Supplemental Results**). Among qPCR positive bloodmeals, 49.1% (52/106) were successfully processed by ddPCR, with 59.6% (31/52) pure wild type, 19.2% (10/52) pure PfKelch13 mutant (n = 3 for C469Y; n = 5 for A675V; n = 2 for both C469Y and A675V), and 23.1% (12/52) having a mixed wild-type and mutant infection (**Figure 4A**). Head and thoraces were processed for all blood-fed (n=330) and non blood-fed (n=299) *Anopheles* mosquitoes. *P. falciparum* sporozoites were detected in 12.6% (41/326) of *An. gambiae s.l.* and 12.5% (38/303) of *An. funestus* mosquitoes. Combining sporozoite-positive mosquitoes from both species with sufficient parasite density signal (n = 34, supplemental methods), 73.5% (25/34) yielded interpretable ddPCR results. Pure wild-type sporozoites were detected in 72.0% (18/25) of sporozoite-positive samples, 16.0% (4/25) were pure C469Y and 12.0% (3/25) were mixed wild-type and mutant infections (n=1 for C469Y; n=1 for A675V, and n=1 for C469Y + A675V; **Figure 4B).**

**Figure 4.**
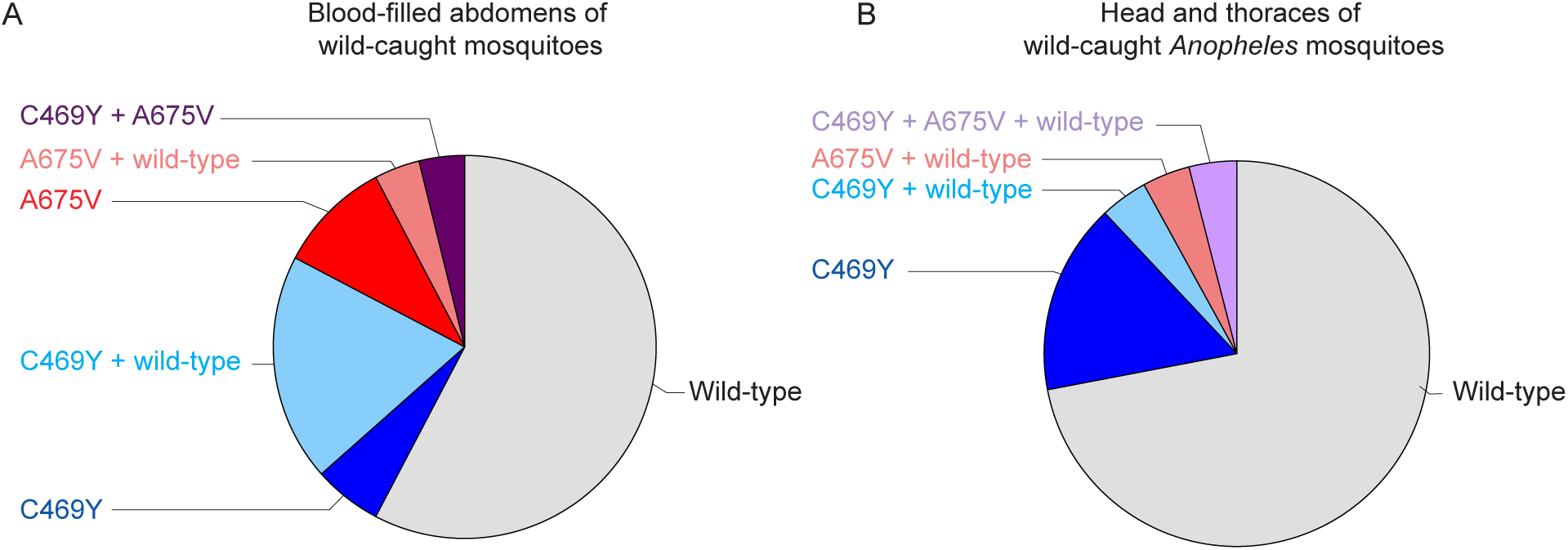
PfKelch13 status of wild-caught mosquitoes. **AB)** Pie charts showing the different categories of PfKelch13 for wild-caught blood-fed mosquito abdomens **(A)** and head-and-thoraces of *Anopheles* mosquitoes **(B)**. PfKelch13 status (wild type, C469Y, A675V) was determined by digital droplet PCR (ddPCR) and is indicated by different colours and in text. Blood-fed mosquito abdomens (A) were selected for ddPCR if sufficient parasite material was detected by 18S qPCR (<35 CT), and included *Anopheles gambiae s.l., Anopheles funestus s.s., Anopheles Zeimanni,* and *Culex spp.* mosquitoes (total N = 52). Sample numbers per category: pure wild type n = 31; pure C469Y n = 3; mixed C469Y n = 10; pure A675V n = 5; mixed A675V n = 2; pure A675V and C469Y n = 2. Head-and-thoraces of *Anopheles* mosquitoes (B) were also selected for ddPCR if sufficient parasite material was detected by 18S qPCR (<35 CT, total N = 25). Sample numbers per category: pure wild type n = 18; pure C469Y n = 4; mixed C469Y n = 1; mixed A675V n = 1; mixed A675V and C469Y (with wild type) n = 1.

## DISCUSSION

We examined gametocyte carriage and transmission potential of parasites with validated markers of ART-R in Uganda. ART-R parasites were present in the majority (65.7%) of patients with uncomplicated malaria, often as multiclonal infections and only detectable by ddPCR. Levels of gametocyte commitment and carriage of mature gametocytes were similar for patients with PfKelch13 mutant or wild-type infections. Using mosquito feeding assays, we observed no indications of differential transmission potential to local *Anopheles* mosquitoes between wild-type and ART-R parasites.

The spread of drug-resistant malaria parasites depends on their ability to produce viable gametocytes. For parasites with resistance to sulfadoxine-pyrimethamine (SP), gametocyte prevalence is increased in the absence and presence of drug pressure (27), and these gametocytes are highly infectious to mosquitoes (28). This contrasts with atovaquone-resistant parasites that suffer very large fitness costs in mosquitoes, probably due to impaired mitochondrial function (29). For ART-R parasites, transmissibility has not been extensively studied. *In vitro* studies with selected (predominantly Asian) parasite lines with partial resistance to artemisinins confirmed their transmission fitness but did not provide strong evidence for increased gametocyte formation or transmission for ART-R parasites (8) and were unavoidably limited by the number of isolates examined. In line with previous studies, we observed gametocytes in 24.0% of patients by microscopy and 43.4% by qRT-PCR (25). Linking these parameters to ART-R was not trivial, considering that approximately 87% of patients presented with multiclonal infections that regularly included both *pfkelch13* wild-type and mutant parasites. Notably, we observed that compared to ddPCR, conventional sequencing greatly underestimated the proportion of infections with ART-R parasites. We did not find evidence for a lower prevalence of *pfkelch13* mutant infections in multiclonal infections, as previously observed for the R622I mutation in Ethiopia (9).

When binning infections as predominantly (or exclusively) wild-type infections, mixed infections, and predominantly or exclusively ART-R infections, we observed no associations with gametocyte commitment or the prevalence or density of mature gametocytes. Our mosquito feeding assays confirmed the transmission fitness of naturally acquired ART-R infections but did not suggest altered mosquito infection rates. Moreover, we observed no evidence that the abundance of ART-R parasites affected the association between gametocyte density and mosquito infection rates, which would have been indicative of an altered transmission efficiency for ART-R parasites.

Lastly, we confirmed the presence of ART-R parasites in wild-caught mosquitoes. For SP resistant parasites, a lower prevalence of parasites with resistance-associated *dhfr* mutations has been observed in wild-caught mosquitoes compared to that in parasites infecting humans (30) providing indirect evidence for a loss in transmission fitness. We observed that 40.4% of mosquito bloodmeals contained ART-R parasites, suggesting a high prevalence of ART-R parasites in the general (largely asymptomatic) population. In sporozoite-positive mosquitoes, ∼28% carried ART-R sporozoites, although interpretation of these data is limited by small numbers and sampling based on convenience outside the seasonal peak in mosquito exposure. Another limitation of our study was that the high complexity of infection, reflective of intense transmission in the study area, may have obscured subtle effects of ART-R on gametocyte dynamics. Gametocyte and transmission dynamics may also differ for PfKelch13 mutations other than C469Y and A675V. Importantly, our study examined gametocyte carriage and transmission potential before treatment was provided. Future studies should examine whether ART-R parasites have, compared to wild-type, elevated post-treatment transmission potential.

In conclusion, we report a high prevalence of parasites with validated markers of ART-R among uncomplicated malaria cases in a highly endemic setting in Uganda. We observed no evidence for enhanced gametocyte formation for ART-R compared to wild-type infections.

## DATA AVAILABILITY

All data are will be deposited in an online repository upon publication.

## ACKNOWLEDGEMENTS

We thank the study team and acknowledge important administrative and technical support from the Infectious Diseases Research Collaboration in Kampala and the Ambrosoli Memorial Hospital in Kalongo, Uganda. We are grateful to the study participants who participated in this study and their families.

