## Supplementary File for "Gametocyte production and infectivity among Ugandan malaria patients infected with *P. falciparum* with partial resistance to artemisinins"

**Supplemental Methods**

**Development of a Multiplex Droplet Digital PCR (ddPCR) for detection of Artemisinin-resistance SNPs**

ddPCR Mutation Detection Assays (BioRad) consisted of one primerset and two different probes that target either the wild-type (WT) sequence or mutant sequence. The mutant ddPCR SNP assay probe was labeled with a FAM-fluorophore, WT SNP assay probe with a HEX-fluorophore. Each ddPCR SNP assay was first set up as stand-alone (singleplex) assay. To use as reference input material, gBlocks with/without the SNP and a total length of 250-280 bp were ordered at IDT. For setting up the ddPCR reaction, the ddPCR Supermix for Probes (No dUTP) was mixed with 1 µL of each WT and Mutant assay and 5 µL of reference material in a total volume of 23 µL as per manufacturer instruction. Droplets were generated with the QX200 Automated Droplet Generator (BioRad), moved to the C1000 Touch Thermal Cycler for thermal cycling (95°C 10 min > 94°C 30 sec, 56°C 60 sec, 39x >98°C 10 min > 12°C constant), followed by readout of droplets on the QX200 Droplet Reader (all BioRad). For data analysis, either the QuantaSoft or QX Manager software was used.

For the first tests, dilution series of gBLocks were prepared in a range of 10^6 to 10^2 copies/ml in MagNAPure elution buffer (Roche) with tRNA carrier (Sigma) at 10 µg/ml to prevent sticking of gBlocks to the tube wall. C469-WT and C469Y or A675-WT and A675V gBlocks were loaded as single targets to check the specificity of each corresponding assay and ensure correct separation of mutant, WT and negative droplet clouds. To verify the assays were giving correct quantification, ddPCR counts were compared to counts of input material. Based on the gBlocks data combined with the data from our actual dataset described in the manuscript, the Limit of Detection (LOD) for both the C469Y and A675V ddPCR SNP assay was determined at 10^2/ml.

As a next step, the influence of matrix on the ddPCR reaction was tested. Negative EDTA whole-blood and microscopy counted wildtype NF54 parasites diluted in EDTA whole-blood in a range of 10^7 to 10^2 parasites/ml were extracted on a MagNAPure 96 automated extractor. For the mutant part of the SNP assays, gBlocks were spiked in the negative blood eluate (as we did not have mutant parasites available in the assay set-u phase ). No effect of the matrix on ddPCR assay performance was observed. Maximum input was also explored and in this experiment 10^7 parasites/ml were still detected without overloading the system (i.e. with sufficient negative droplets to meet manufacturer recommendations). Loading a high quantity of WT parasites (10^7 parasites/ml) together with low amounts of mutant gBLock in the correct matrix also did not alter the LOD of both C469Y and A675V mutant assay. Next to this, both the C469Y and A675V SNP assay were used to count the same NF54 WT parasite material. Estimates of each individual assay were compared and showed near identical results, confirming once more the independent assays are achieving the correct quantification of their target sequence.

Next, it was tested if the C469Y mutant probe would also recognize the C469F SNP but with an altered efficiency due to the one mismatch in the probe, hence changing the positioning of the mutant cloud. This was indeed the case, and this assay was from there on used as C469Y/F SNP assay.

A temperature gradient from 55 to 60 degrees was run to determine the optimal annealing/elongation temperature that optimizes separation between positive and negative droplets while minimizing rain. For both assays, higher temperature resulted in a trend of mutant and WT clouds appearing lower towards the negative, so the range of separation became smaller. At 60 degrees the mutant and WT clouds could no longer be separated from the negative cloud. Annealing/elongation temp was therefore set at 56 degrees.

In a next step, we combined the C469Y/F and A675V SNP assays. The QX200 reader has 2 channels available (FAM/HEX) and each SNP assay was run as mutant-FAM and WT-HEX. For multiplexing, the assay probes were adjusted to the following setup: C469Y/F-FAM, C469-WT-HEX, A675V-HEX and a A675-WT-DARK-Cy5.5 probe to block off the A675-WT assay part. We optimized the amount of each assay in the multiplex ddPCR reaction to maximize separation between each different mutant or WT cloud and the negative cloud. The selected (optimal) amounts were 0.5 ul of C469Y/F-FAM probe, 0.4 ul of C469-WT-HEX probe, 1 ul of the A675V-HEX and A675V-WT-DARK-Cy5.5 probe. The MPX ddPCR was used to screen for the presence of mutations in our samples (**Figure 1**). In the FAM channel, the double positive FAM cloud (both C469Y and C469F mutant target present in 1 droplet) does not separate out from the C469Y cloud. In case we detected both mutant sequences present in a sample, this sample needed to be run in separate C469Y and C469F SNP assays for correct quantification. As we were using the C469-WT assay to determine WT counts, all A675V mutant parasites will also give a signal in this WT assay. To resolve this issue, if a sample was determined positive for A675V it was repeated in the single A675V SNP assay for accurate quantification (**Figure 1**). For calculation of mutant vs WT percentages,the C469Y/F counts from the MPX ddPCR were combined with the A675V single assay counts of WT and A675V mutant. If no A675V was detected in the MPX assay, counts from the MPX ddPCR were used for calculation of percentages.

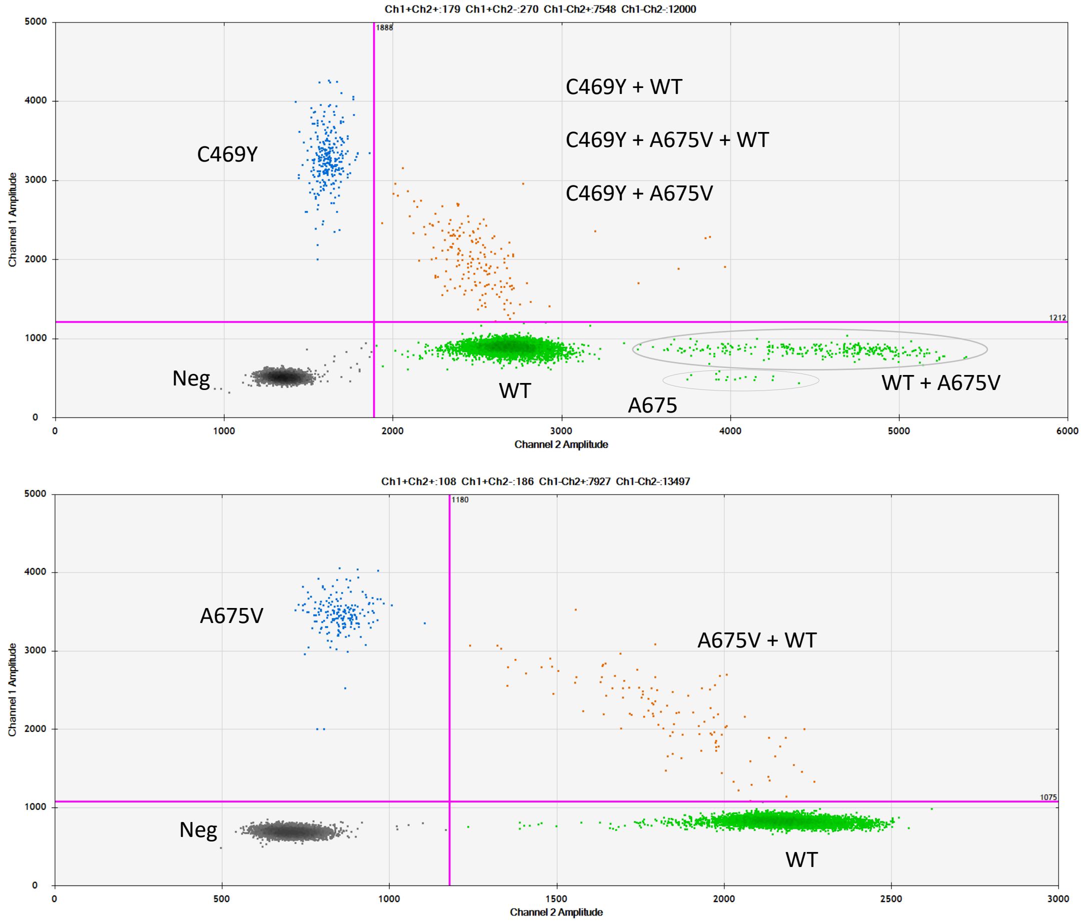


***Figure 1**. Droplet populations for the multiplex (upper panel) and A675V single plex (lower panel) assay of a single sample that was positive for C469Y, A675V, and wild-type. The sample was first run in multiplex and then repeated in singleplex for correction quantification of both A675V and wild-type parasites. Population types are written in text.*

**Defining the success rate of ddPCR in relation to parasite density in field-caught *Anopheles* mosquitoes**

To assess the presence of mutant parasites in field-caught mosquitoes, we used the ddPCR MPX assay for i) blood-fed abdomen samples, and ii) head and thorax samples. 18S qPCR was done to determine the parasite density of these samples. An initial pilot with a single-plex ddPCR (C469Y) was performed using eight parasite positive samples (determined by 18S qPCR [1]) in each group (16 samples in total). This was done to determine the sensitivity of the ddPCR on these sample types (**Table 1**). All eight blood meal pilot samples had an 18S qPCR CT value below 32 and mutant (C469Y) or wild-type parasites were detected in all samples. The 18S qPCR CT values were higher in head and thorax samples; all samples with an CT-value above 35 did not result in a ddPCR signal. For this reason, blood-fed abdomens and head and thorax samples were selected for ddPCR to determine p*fkelch13* mutations if the 18S qPCR signal was < 35 CT; samples with higher CT-values were not processed and considered indeterminate.

| | **18S CT value** | **ddPCR counts** | |
|---|---|---|---|
| | | C469Y | WT |
| **Blood meal** | | Par/ml | Par/ml |
| Sample 1 | 31.15 | 0 | 4922 |
| Sample 2 | 24.64 | 199180 | 52440 |
| Sample 3 | 29.16 | 4600 | 1104 |
| Sample 4 | 29.81 | 0 | 6900 |
| Sample 5 | 28.11 | 736 | 27140 |
| Sample 6 | 28.61 | 0 | 12420 |
| Sample 7 | 29.12 | 6900 | 2530 |
| Sample 8 | 29.75 | 0 | 8740 |
| **Head and thorax sample** | | | |
| Sample 9 | 35.45 | 0 | 0 |
| Sample 10 | 30.81 | 5367 | 0 |
| Sample 11 | 37.06 | 0 | 0 |
| Sample 12 | 36.26 | 0 | 0 |
| Sample 13 | 35.14 | 0 | 0 |
| Sample 14 | 31.88 | 422 | 1648 |
| Sample 15 | 35.14 | 0 | 192 |
| Sample 16 | 36.95 | 0 | 0 |

*Table 1. ddPCR pilot set of mosquito bloodmeal and head and thorax samples.*

**Supplemental Results**

| | Parasite prevalence (18S qPCR) in blood fed mosquito abdomens (n/N, %) |
|---|---|
| Total | 106/269 (39.4) |
| *An. funestus* s.s | 19/37 (51.4) |
| *An. gambiae* s.l. | 19/23 (82.6) |
| *Culex spp.* | 68/209 (32.5) |

**Supplementary Table 1:** Parasite prevalence in blood-fed wild caught mosquito abdomens.

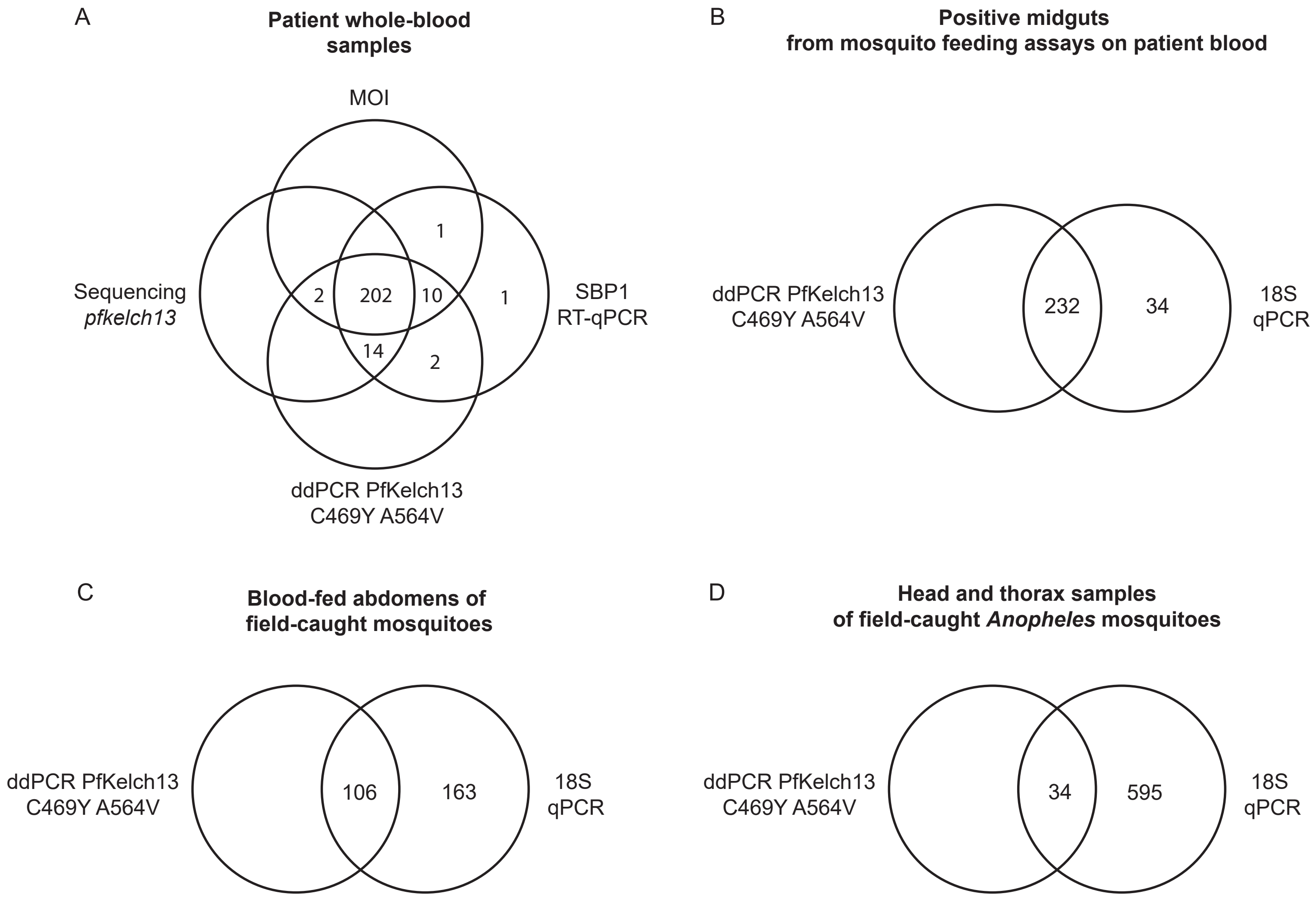


**Supplementary Figure 1**. Laboratory procedures and sample numbers per sample type. Venn diagram showing the (overlap in) molecular assays for a) patient whole-blood samples, b) mosquito midguts with at least one oocyst (positive midguts) dissected after a membrane feeding assay, c) abdomens of field-caught and blood-fed mosquitoes, and d) head-and-thoraces of field-caught Anopheles mosquitoes. A) For patient whole-blood samples, the assays included RT-qPCR on SBP1 transcripts to determine ring-stage parasite densities, multiplicity of infection (MOI) determination through MAD4HatTeR, sequencing of PfK13, and a digital-droplet-PCR (ddPCR) to determine the quantity of PfK13 C469Y, A675V and wild-type (WT) parasites. A total of 235 patient samples were collected; 3 samples were negative for all four assays and not included in the figure. B) For positive midguts from membrane feeding assays, 18S qPCR was initially performed and 18S positive samples were selected for ddPCR. A total of 324 positive midguts were selected for 18S quantification, 57 had no 18S signal and was not included in ddPCR, 1 sample was not included in both assays. 34 samples with 18S signal were negative in ddPCR. C) A total of 269 field-caught blood-fed mosquito abdomens were included in 18S qPCR; 106 samples with sufficient parasite material were selected for ddPCR. Of these, 53 samples had an interpretable ddPCR result. D) A total of 629 head and thorax samples from field-caught Anopheles mosquitoes were selected for parasite quantification by 18S qPCR; 34 samples with sufficient parasite material were selected for ddPCR. Of these, ddPCR resulted in an interpretable signal in 25 samples. ABCD) An assay was considered negative when it resulted in a signal of zero (for SBP1 and 18S) or when the assay was not performed.

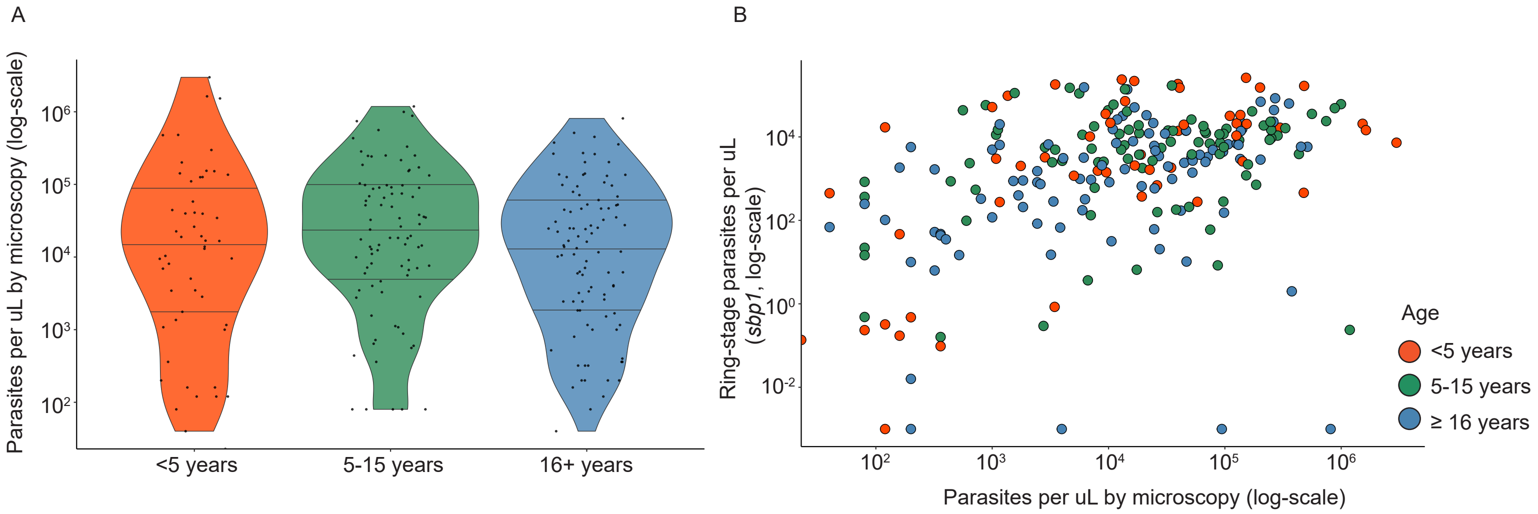


**Supplementary Figure 2**. Parasite density in relation to participant age. A) Violin plot of parasite density measured by microscopy for the three different age groups (<5 years old, 5-15 years old, 16+ years old). B) Scatter plot showing the relation between parasite densities measured by micrscopy versus the ring-stage density measured by RT-qPCR targeting *sbp1* transcript levels in the same patient. Colours indicate the participant's age (in categories).

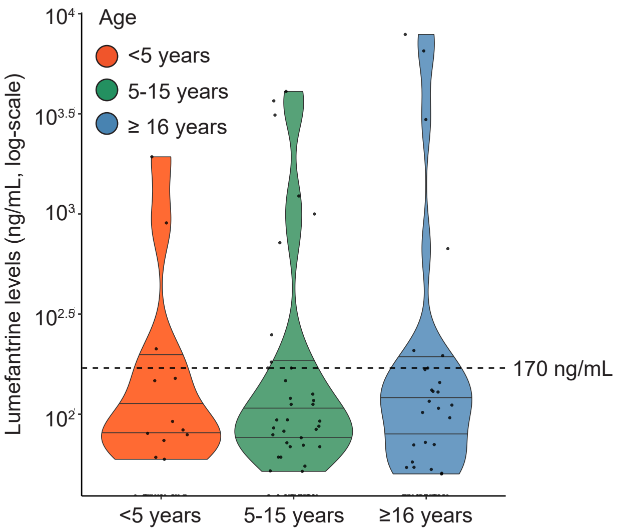


**Supplementary Figure 3**. Lumefantrine plasma levels in relation to participant age. Violin plot showing lumefantrine levels, dashed line indicates the lumefantrine level corresponding to having received treatment in the past three weeks.

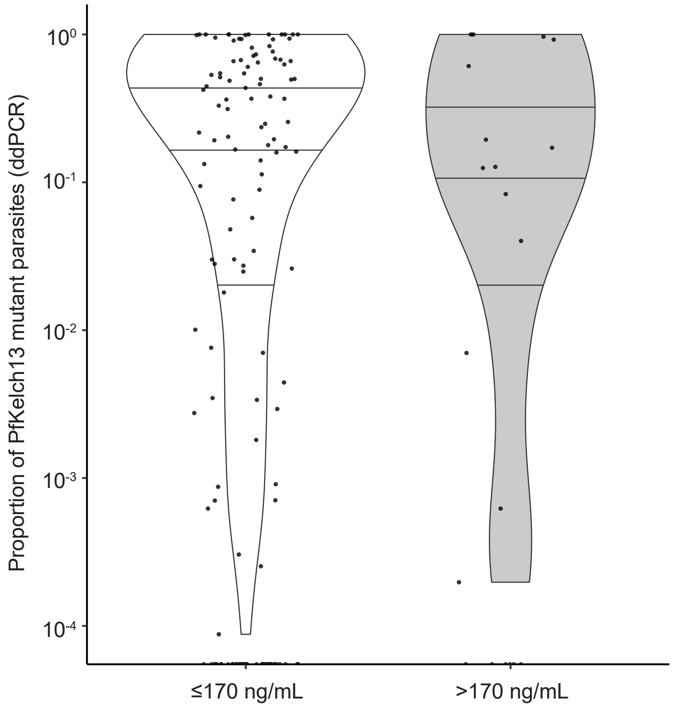


**Supplementary Figure 4**. Proportion of PfKelch13 mutant parasites in relation to plasma lumefantrine levels. Violin plot showing different proportions of PfKelch13 mutant parasites in patient whole-blood. PfKelch13 mutant parasites were detected using digital droplet PCR (ddPCR) at the 469 and 675 locus. Lumefantrine levels were categorized based on the lumefantrine level corresponding to having received treatment in the past three weeks (170ng/mL). The difference in abundance of PfKelch13 mutant parasites between the two lumefantrine categories was not statistically significant ($p = 0.701$).

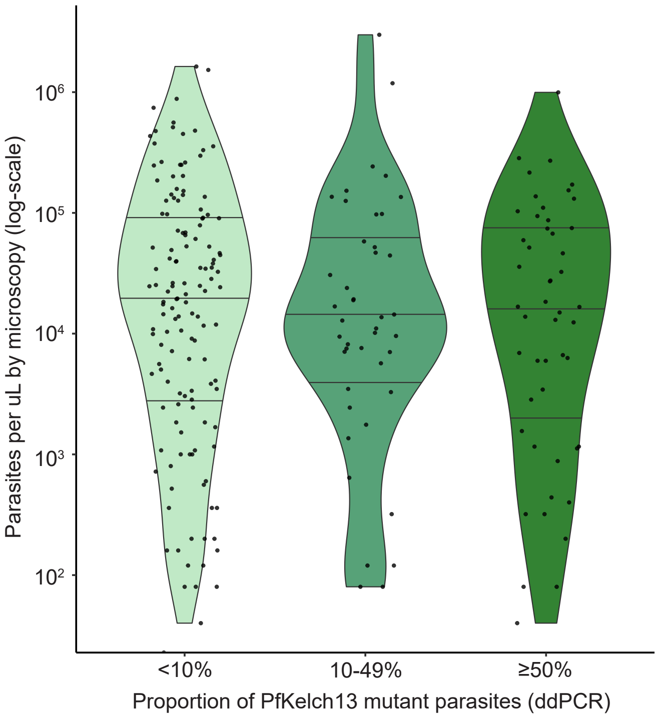


**Supplementary Figure 5.** Parasite density in relation to the proportion of PfKelch13 mutant parasites. Violin plot showing parasite densities measured by microscopy for three different categories based on the proportion of PfKelch13 mutant parasites. PfKelch13 mutant parasites were detected using digital droplet PCR (ddPCR) at the 469 and 675 locus.

Digital droplet PCR (ddPCR)

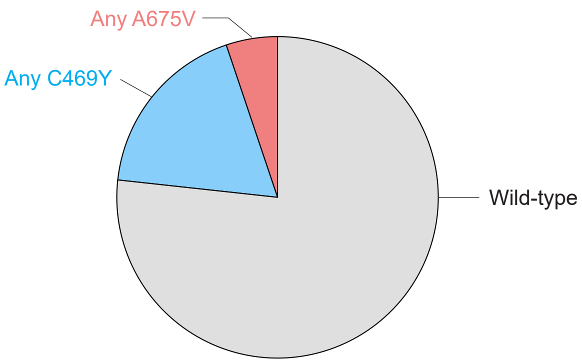


**Supplementary Figure 6**. PfKelch13 mutant status in mosquito midguts after feeding on patient blood. PfKelch13 mutant parasites were detected using digital droplet PCR (ddPCR) at the 469 and 675 locus. Sample numbers per category: pure wild-type (n=178), any C469Y present (n=42, of which 7 pure C469Y), any A675V present (n=12, of which 8 pure A675V).
